# Booster vaccination protection against SARS-CoV-2 infections in young adults during an Omicron BA.1-predominant period: a retrospective cohort study

**DOI:** 10.1101/2022.03.08.22272056

**Authors:** Jiayue Wan, Casey L. Cazer, Marin E. Clarkberg, Shane G. Henderson, Scarlett E. Lee, Genevive Meredith, Marwan Osman, David B. Shmoys, Peter I. Frazier

## Abstract

**Background:** While booster vaccinations clearly reduce the risk of severe COVID-19 and death, the impact of boosters on SARS-CoV-2 infection has not been fully characterized: doing so requires understanding their impact on asymptomatic and mildly symptomatic infections that often go unreported but nevertheless play an important role in spreading SARS-CoV-2. We sought to estimate the impact of COVID-19 booster doses on SARS-CoV-2 infection in a vaccinated population of young adults during an Omicron BA.1-predominant period.

**Methods and Findings:** We implemented a cohort study of young adults in a college environment (Cornell University’s Ithaca campus) from a period when Omicron BA.1 was the predominant SARS-CoV-2 variant on campus (December 5 to December 31, 2021). Participants included 15,800 university students who completed an initial vaccination series with a vaccine approved by the World Health Organization for emergency use, were enrolled in mandatory at-least-weekly surveillance PCR testing, and had no positive SARS-CoV-2 PCR test within 90 days before the start of the study period. Robust multivariable Poisson regression with the main outcome of a positive SARS-CoV-2 PCR test was performed to compare those who completed their initial vaccination series and a booster dose to those without a booster dose.

1,926 unique SARS-CoV-2 infections were identified in the study population. Controlling for sex, student group membership, date of completion of initial vaccination series, initial vaccine type, and temporal effect during the study period, our analysis estimates that receiving a booster dose further reduces the rate of having a PCR-detected SARS-CoV-2 infection relative to an initial vaccination series by 56% (95% confidence interval [42%, 67%], P <0.001). While most individuals had recent booster administration before or during the study period (a limitation of our study), this result is robust to the assumed delay over which a booster dose becomes effective (varied from 1 day to 14 days). The mandatory active surveillance approach used in this study, under which 86% of the person-days in the study occurred, reduces the likelihood of outcome mis-classification. Key limitations of our methodology are that we did not have an *a priori* protocol or statistical analysis plan because the analysis was initially done for institutional research purposes, and some analysis choices were made after observing the data.

**Conclusions:** We observed that boosters are effective, relative to completion of initial vaccination series, in further reducing the rate of SARS-CoV-2 infections in a college student population during a period when Omicron BA.1 was predominant; booster vaccinations for this age group may play an important role in reducing incidence of COVID-19.

## Introduction

COVID-19 vaccines reduce SARS-CoV-2 infection and symptom severity [1], yet breakthrough infections occur, especially with the Omicron (B.1.1.529) variant [2, 3]. The effectiveness of Food and Drug Administration (FDA)-authorized or approved vaccines BNT162b2, mRNA-1273, and Ad26.COV2.S in preventing SARS-CoV-2 infection has dropped dramatically due to immune evasion and waning of vaccine-induced immunity over time [4-7]. The Omicron variant exhibits immune-system escape as the result of several mutations [8, 9]; this, and the high transmissibility of the Omicron variant, are leading to higher infection rates, strain on healthcare systems, and increased mortality [10, 11]. With the emergence of new variants combined with waning immunity, the CDC recommends a booster six months after an initial mRNA vaccine series or two months after Ad26.COV2.S vaccination to prevent symptomatic and severe outcomes of COVID-19 [7, 12]. The booster dose elicits an increase in antibody neutralization titers against the Omicron variant, and causes affinity maturation leading to a better antibody response, maintaining long-term protection against severe COVID-19 outcomes [12-15].

While boosters are understood to be effective against severe disease, hospitalization [16] and symptomatic infections [12, 17] resulting from the Omicron variant, limited information is available about their effectiveness against asymptomatic and mild symptomatic infections that may go unreported in the absence of asymptomatic surveillance testing. Asymptomatic and mild symptomatic infections play an important role in spreading SARS-CoV-2 [18]. Moreover, the clinical course of COVID-19 is understood to vary by age [19]; current estimates of booster effectiveness based on the general population may not apply to cohorts whose age distribution differs substantially from that of the general population.

Leveraging PCR testing data derived from mandatory SARS-CoV-2 surveillance program through December 14, 2021 for the study population and required departure tests before students leaving the Ithaca campus, we sought to estimate the effectiveness of COVID-19 boosters in reducing SARS-CoV-2 infections in a vaccinated population of students at Cornell University’s Ithaca campus during December 5 to December 31, 2021, when there was an explosive increase in Omicron cases to the point where the Omicron variant (BA.1) became predominant [20] and 1,926 SARS-CoV-2 infections were identified.

## Methods

We used a retrospective cohort study to estimate the effectiveness of COVID-19 boosters in reducing SARS-CoV-2 infections. Specific to our context was a highly vaccinated study population, and a time period where Omicron BA.1 was the dominant viral strain. We did not have an *a priori* protocol or statistical analysis plan. As detailed in **Supplementary Section S1**, our initial analysis [21] came as part of Cornell University’s institutional response to COVID-19 during an Omicron-driven outbreak among a highly vaccinated population. This plan was made after observing aggregate infection rates that compared those who had uploaded early proof of booster vaccination (less than 17% of those in the study population who would eventually provide documentation) against those who had not. We subsequently developed a study plan that modified two of the covariates in this analysis (adding sex and changing how primary vaccine series completion date was coded) as we hypothesized that they are confounders. We then made additional modifications in response to reviewer requests. As described in **Supplementary Section S1**, estimates of vaccine booster effectiveness were not sensitive to these changes in the analysis.

We utilized de-identified student data from Cornell University’s COVID-19 surveillance database. We included polymerase chain reaction (PCR) positive cases identified at Cornell surveillance testing sites, the campus student health service, and a sampling site operated by the local hospital system during the period of December 5, 2021 to December 31, 2021 (referred to as the “Omicron outbreak” or the study period), in which the Omicron variant (BA.1) was the predominant strain. Samples collected at Cornell surveillance sites and the local hospital’s sampling site were tested using EZ-SARS-CoV-2 Real-Time RT-PCR [22] within a two-stage Dorfman procedure [23] using pools of size 5. The assay targets two highly-conserved regions of the SARS-CoV-2 N gene and the Omicron variant is not expected to significantly affect test performance. Samples collected at the campus student health service were tested using the Cepheid Xpert Xpress SARS-CoV-2 Assay [24]. The FDA reports that the introduction of the Omicron variant does not appear to have resulted in a significant change in the test performance of this assay [25]. This work was completed as a part of Cornell’s institutional planning and preparedness, designated as exempt from IRB review by the Cornell IRB.

### Study Context

In July 2020, as a part of its reopening strategy, Cornell implemented a robust COVID-19 surveillance testing program for students, faculty, and staff (1,637,394 PCR tests performed as of February 2, 2022). At the start of December 2021, 23,389 students were enrolled in full-time academic programs at Cornell’s Ithaca campus. Masks were required in all on-campus buildings. COVID-19 vaccination was required for all students (97% vaccinated), though medical or religious exemptions were granted. Boosters were encouraged (October 21, 2021) before being required (January 31, 2022). Through December 14, 2021, all undergraduates and professional (veterinary, business, and law) students (*n* = 17,017) were enrolled in at-least-weekly mandatory PCR surveillance (compliance: 99.8% for surveillance tests scheduled between November 29, 2021 and December 14, 2021) using an anterior nasal swab sample. 86% of person-days included in the study (see the **Study Population** section below) fell within this period. In addition, students leaving the Ithaca campus before the end of the study period (82% of students in the study population indicated that they left before the end of the study period through a Cornell-managed checklist) were required to seek an additional test shortly before departure. Additionally, students were prompted to seek a PCR test if they were identified as a close contact of a case or if they developed COVID-like symptoms. Free PCR testing was available to any constituent of the university 7 days per week. Whole genome sequencing confirmed the presence of the Omicron variant (BA.1) in Cornell community COVID-19 surveillance samples collected on December 1, 2021; by December 11, 2021, Omicron was the predominant variant on the Ithaca campus [20].

### Study Population

Study inclusion criteria were: being an active student at Cornell’s Ithaca campus enrolled in mandatory surveillance testing, being vaccinated with a vaccine approved by the World Health Organization (WHO) for emergency use at the time of the study (two doses of BNT162b2, two doses of mRNA-1273, one dose of Ad26.COV2.S, or two doses of other WHO-approved vaccines), and having no positive SARS-CoV-2 PCR test within 90 days before the start of the study period (**Figure 1**). Students were excluded from the study population if they were not subject to mandatory surveillance testing (i.e., graduate students) (*n* = 6,372), tested PCR-positive within 90 days before the start of the study period (*n* = 214), did not have at least one PCR surveillance test between November 5, 2021 and December 4, 2021 (*n* = 130), had not completed a WHO-approved initial vaccine series (*n* = 701), had an unspecified sex (*n* = 9), or had invalid vaccination records (*n* = 163). Our study included 15,800 Cornell students. Students with no PCR test records during the study period (*n* = 285) were considered lost to follow up. Students were required to upload a photo of a vaccine card documenting their booster status via an online form or request an exemption by February 1, 2022 [26]. According to vaccination records that were uploaded and validated by February 8, 2022, 11.9% of the study population (*n* = 1,876) received a booster before the study period, with the earliest booster dose administered on August 3, 2021. Our analysis does not distinguish the booster vaccine manufacturer, although this information is available in **Supplementary Table S2**.

**Figure 1:**
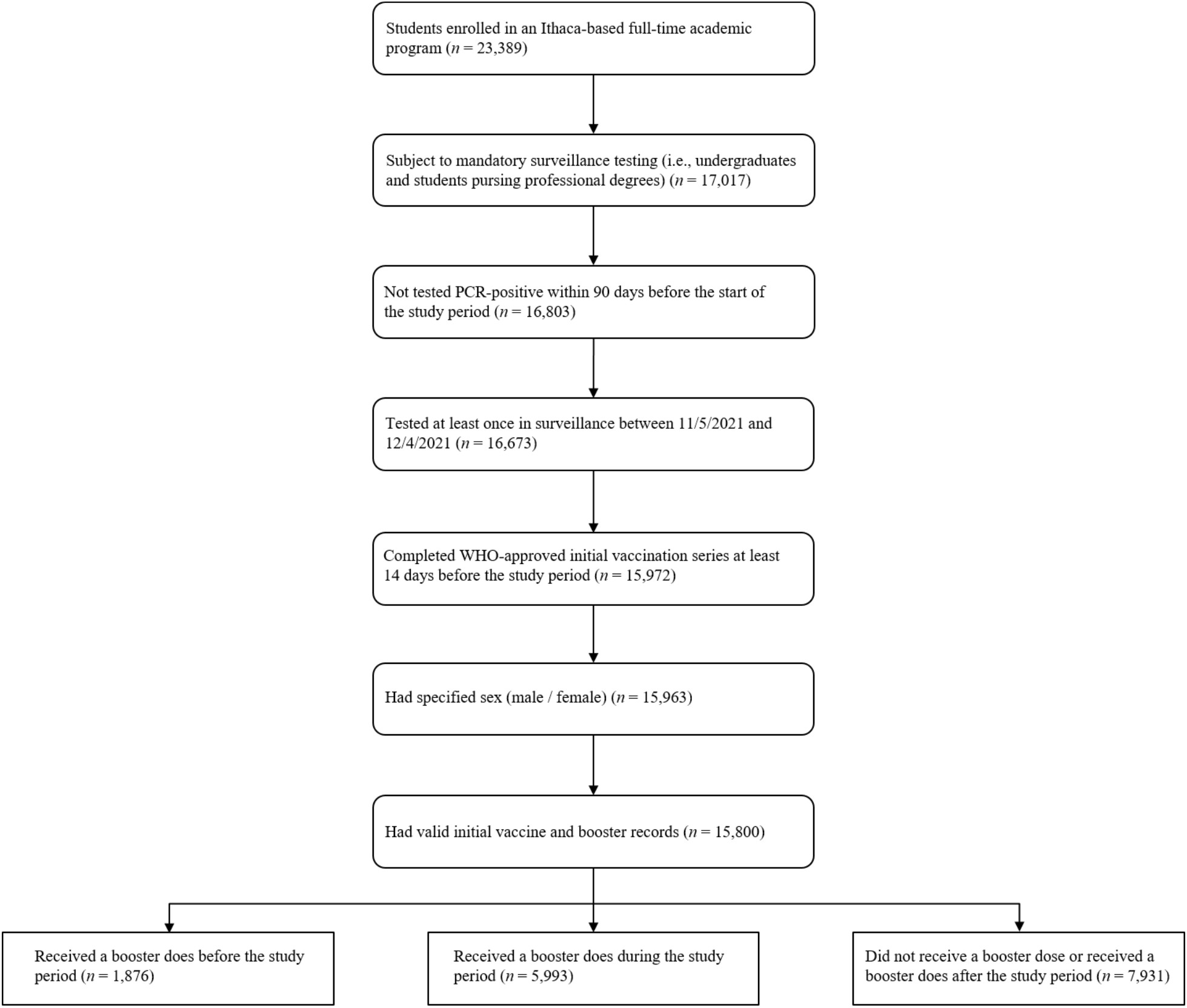
Enrollment of the study population. Students were included if they were enrolled in an full-time academic program based at Cornell’s Ithaca campus in Fall 2021, were subject to mandatory surveillance testing, had no positive SARS-CoV-2 PCR test within 90 days before the start of the study period, had at least one PCR test between November 5, 2021 and December 4, 2021, had a valid vaccination record indicating completion of WHO-approved initial vaccination series at least 14 days before the study period, and specified their sex in university records.

### Statistical Analysis

The primary outcome of our study was PCR-confirmed SARS-CoV-2 infection. To estimate the effectiveness of COVID-19 boosters against SARS-CoV-2 infections during this Omicron-predominant period, we calculated the person-days that each student contributed to the boosted and non-boosted population during the study period. The total number of person-days contributed by a student is the number of days between December 5, 2021 and either their final test date in the study period or their first PCR-positive test date, whichever comes first. Of the study population, 38,023 PCR tests were performed during the study period. 98.2% of the students (*n* = 15,515) had at least one PCR test (mean = 2.4 tests, median = 2.0 tests, SD = 1.2 tests), hence contributing at least one person-day (**Supplementary Figure S1**). We assume that booster vaccinations become effective 7 days after administration, based on results from a study of the Israeli general population [27]. As a result, each person-day has its booster status labeled as boosted (1) or control (0) based on whether the associated individual received their booster dose at least 7 days before that day (**Supplementary Figure S2**).

We performed a multivariable Poisson regression to estimate the effect of receiving a booster dose on having a positive PCR-based diagnosis during the study period. The unit of observation is an exposure period of an individual (measured in person days). An offset equal to log(person-days), i.e., the natural logarithm of the number of days in the exposure period, is added in the regression model for each exposure period. Each exposure period lasts until the end of the week (Week 0 is December 5 to December 11, etc.), a change in booster status (in which case a week is split into two exposure periods), or the person completes their last test in the study, whichever comes first. This model can be seen as a Cox proportional hazards analysis [28], in which it is assumed that exposure risk accrues at a constant rate within each exposure period. To account for possible within-individual correlation among observations over time (i.e., the risk of some individual contracting the disease is likely persistent over time due to individual behavior), we used generalized estimating equations (GEE) [29, 30] with an exchangeable correlation structure over weeks in the study period to obtain consistent coefficient estimates and robust variance estimates for the regression model.

We controlled for sex, student group (undergraduate or professional student), fraternity/sorority participation (yes or no), varsity athletic team participation (yes or no), initial vaccine type (BNT162b2, mRNA-1273, Ad26.COV2.S, or other WHO-approved vaccines), initial vaccination series completion date (date of receiving 1 dose of Ad26.COV2.S or 2 doses of other WHO-approved vaccines, grouped by months from January 2021 to November 2021), and temporal effect during the study period (grouped by week). We also created univariable models for each of these covariates to obtain unadjusted incidence rate ratios. We included student group, fraternity/sorority participation, and athletic team participation as covariates because case investigation of data from before the study period suggested that these covariates explained much of the heterogeneity in the risk of infection across students [31]. Undergraduate students were divided into 3 subgroups based on fraternity/sorority and athletic team participation. Students who were in both fraternities/sororities and athletic teams were classified in the fraternity/sorority group. Thus, we had six student groups: (1) undergraduate fraternity/sorority participants; (2) undergraduate athletes not in Group 1; (3) undergraduates not in Groups 1 or 2; (4) law students; (5) business graduate students; (6) veterinary students. Initial vaccination series completion date was included as a covariate to adjust for heterogeneous social behavior, inclination to receive vaccination, and waning vaccine immune response. The week during the study period was included as a categorical covariate to control for time-varying prevalence and the resulting risk of exposure. This mitigates bias that would have otherwise been created by temporal variation in the fraction of on-campus students boosted. We used a categorical covariate rather than a continuous one because prevalence did not change linearly with time. We did not include age as a covariate because there is little age variation in the study population (**Supplementary Figure S3**). The dependent variable was whether a student tests PCR-positive for COVID-19 in a particular exposure period. The regression model was:

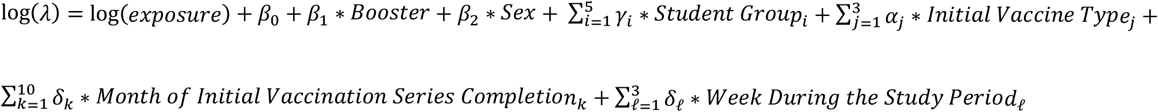

where *λ* is the incidence rate, and greek letters on the right hand side of the specification are the regression coefficients. The *p-*values for the estimated coefficients and the 95% confidence intervals for the adjusted incidence rate ratios were adjusted using the Bonferroni correction [32].

To estimate the effectiveness of a booster dose we used

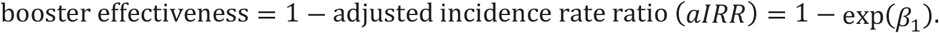

We performed several robustness checks. To assess the effect of students leaving campus at the end of the semester and the accordingly paused mandatory surveillance, we performed a secondary GEE Poisson regression with the same specification as above, but for a shorter study period (from December 5, 2021 to December 18, 2021, the last day of exams).

To assess the possibility of bias remaining in our analysis despite the included controls (e.g., bias due to days since booster dose administration), we performed a GEE Poisson regression with multiple classes for the booster status (unboosted, 0-6 days after booster administration, ≥ 7 days after booster dose administration). We hypothesized that booster vaccination has negligible effect 0-6 days after administration.

We further performed a sensitivity analysis on the delay to boosted status, allowing person-days to count as boosted only after this delay after administration had elapsed, varying this delay from the day after booster administration (day 1) to 14 days (**Supplementary Figure S2**). We emphasize that varying this parameter only affects the booster status of person days from people that were boosted within or shortly before the start of the study period. We hypothesized that the shorter delays would result in smaller but still statistically significant booster effectiveness estimates as person-days from 0-6 days after booster administration would dilute the effect observed in the ≥ 7 days after booster administration group. We expected to see similar estimates of booster effectiveness for all delays ≥ 7 days. We emphasize that this analysis is not designed to comment on how quickly boosters become effective, but instead only on the robustness of our main findings to assumptions about this delay parameter.

We also performed a logistic regression (with GEE) to assess the robustness of our conclusions to model specification. The unit of observation is a person-day. We assumed that booster vaccinations become effective 7 days after administration. The dependent variable is whether a student tests PCR-positive for SARS-CoV-2 on a particular day. The regression model was:

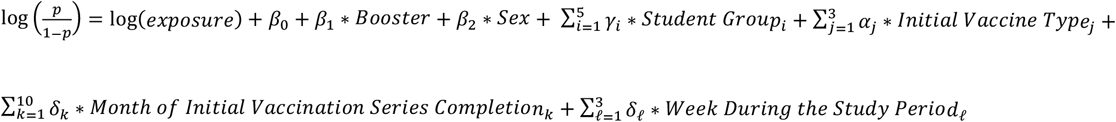

where *p* is the probability of infection, and greek letters on the right hand side of the specification are the regression coefficients. This logistic model can be seen as a form of discrete-time interval-censored survival model, in which a person’s endpoint is the first day on which they tested positive and the right-censoring date is the date of their last negative test. As with the Poisson regression model, information bias is possible because testing did not occur every day. We approximated the incidence rate ratio using the adjusted odds ratio (aOR) when estimating the booster effectiveness.

A subset analysis of undergraduate students living on campus was performed to assess the robustness of our results to additional kinds of student clustering (i.e., residence) through which virus could spread. The description of this analysis is included in **Supplementary Section S2**.

All statistical analyses were performed in Python (V3.7.11), using the statsmodels package (V0.13.2). All code is publicly available (https://github.com/jiayuewan/booster_effectiveness). The data is protected by an institutional data requirement at Cornell that covers the use of data collected during COVID-19 surveillance and is maintained by Cornell University’s Office of the Vice President for Research and Innovation. It can be requested from that entity.

## Results

Boosted students were more likely to be female, professional students, and have an earlier initial vaccine series than unboosted individuals (**Table 1**). During the study period, a total of 1,926 PCR-positive cases were identified and reported out of 15,800 students included in this analysis (overall infection risk of 12.2%). None of the cases required hospitalization. The overall infection risk among students that received a booster dose before December 5, 2021 is 6.2% (117 PCR-positives out of 1,876 boosted students), whereas the infection risk among students that did not receive a booster before December 5, 2021 is 13.0% (1,809 PCR-positives out of 13,924 students). The cumulative incidence among students who received a booster before December 5, 2021 was approximately half the cumulative incidence among students who were unboosted at that time (**Figure 2**). Similarly, the incidence rate was 0.6 per 100 person-days (127 PCR-positive cases/19,842 person-days) among students who had received the booster dose ≥ 7 days earlier, compared to 1.3 per 100 person-days (1,799 PCR-positive cases/135,214 person-days) among those who had not received the booster dose or had received the booster dose < 7 days earlier.

**Table 1:**
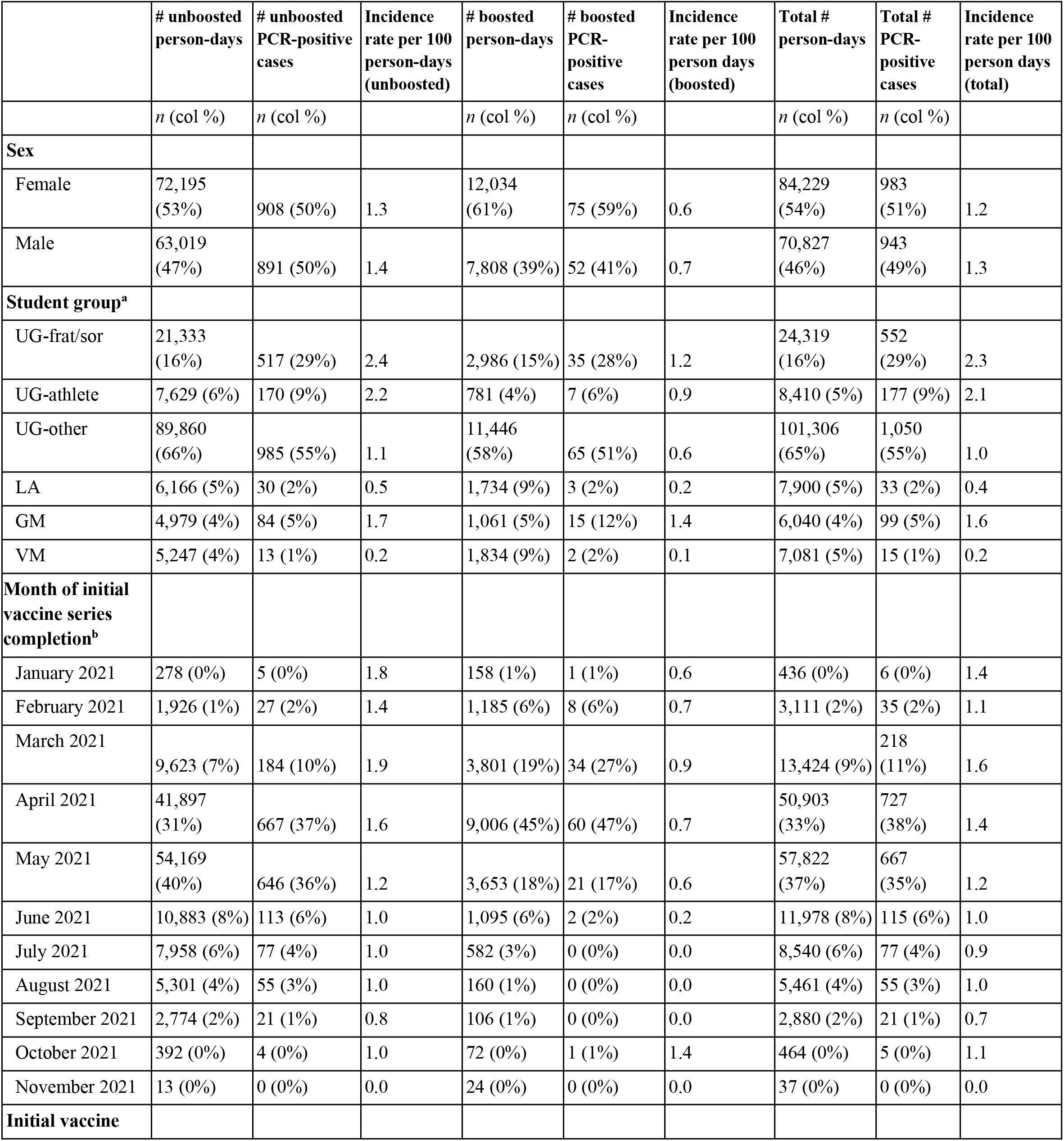
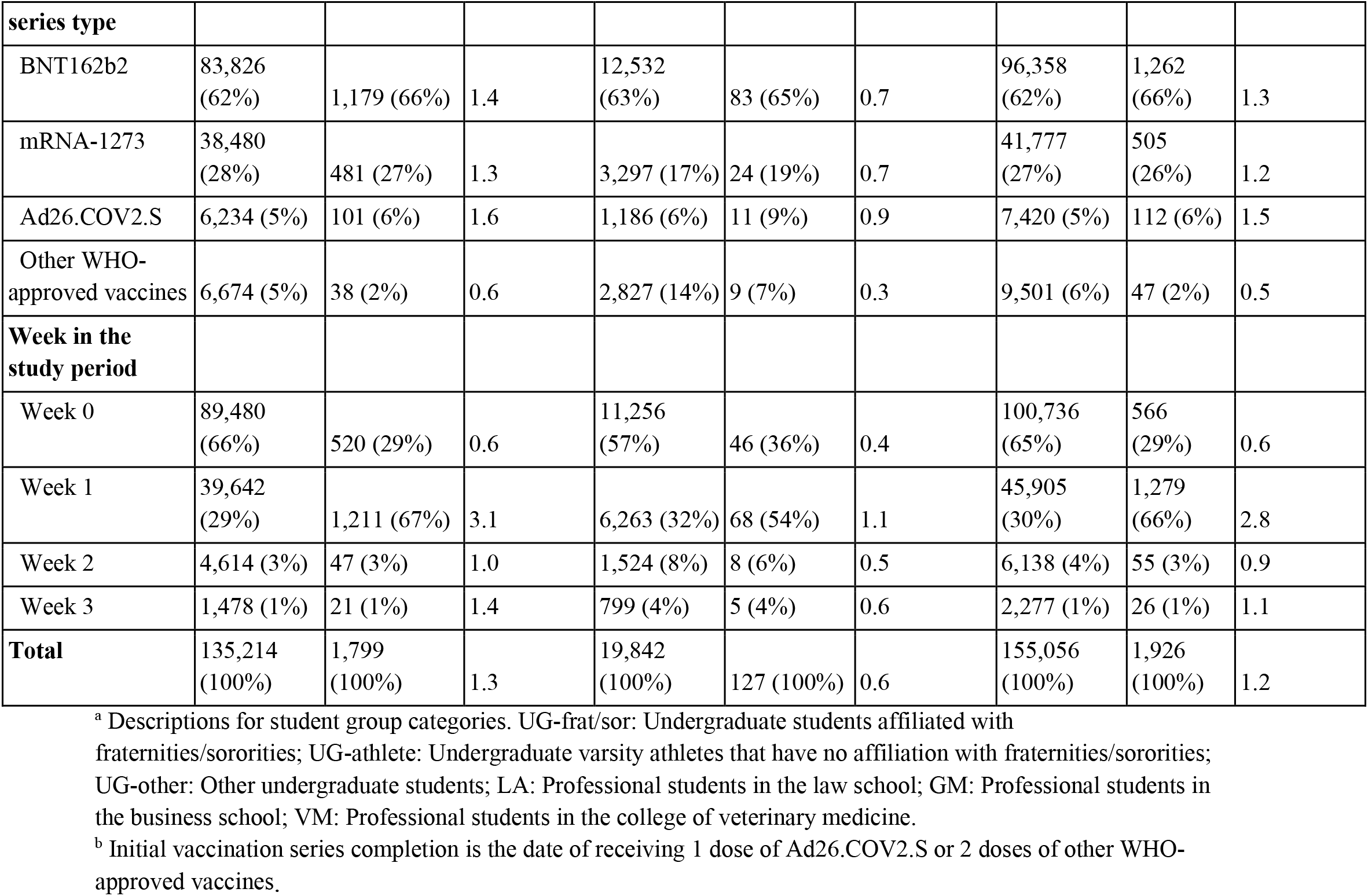
Distribution of person-day data by sex, student group, initial vaccination series completion date, initial vaccine type, and week in the study period.

**Figure 2:**
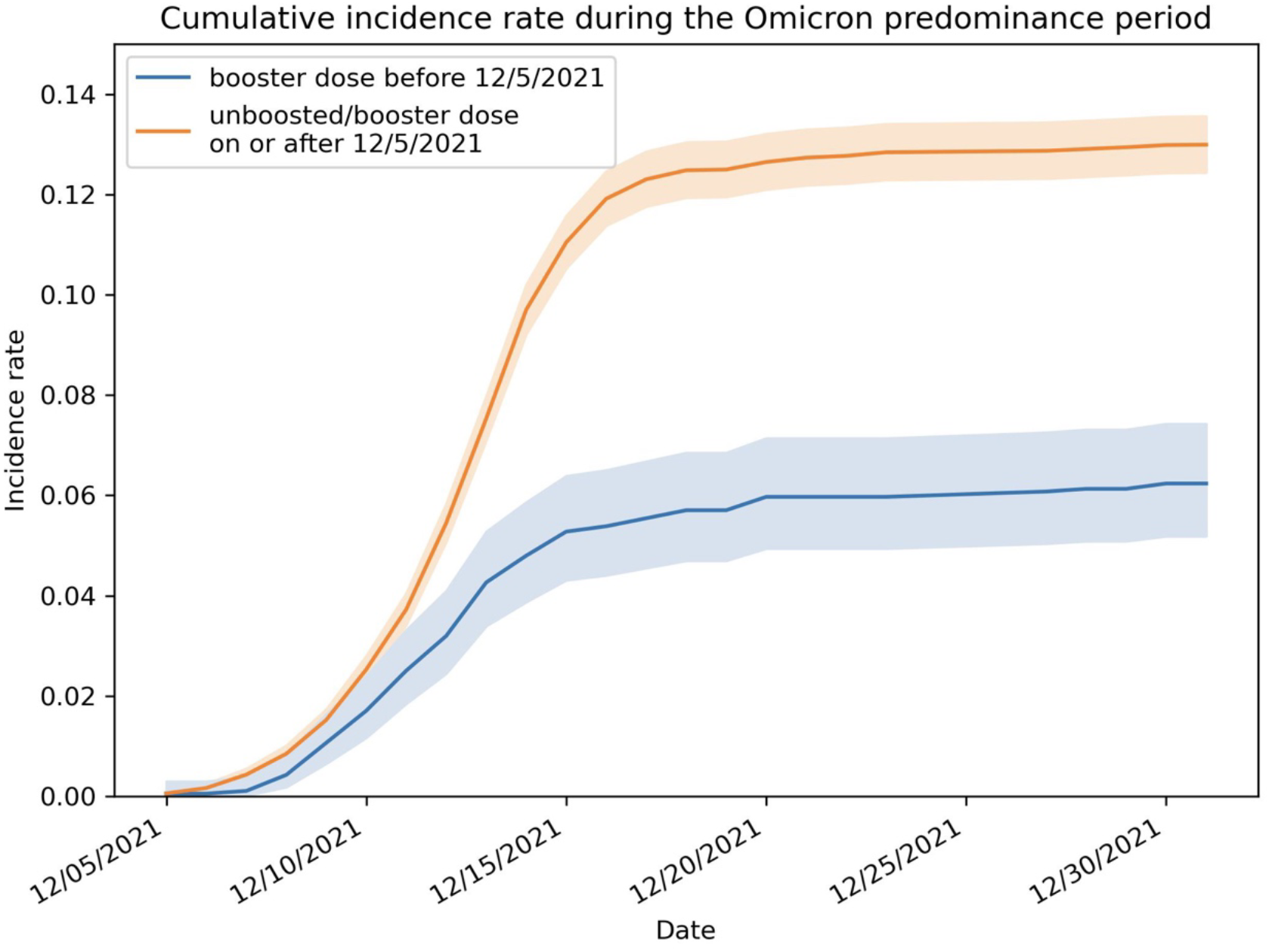
SARS-CoV-2 infection cumulative incidence rate (number of infections per person) and its 95% confidence interval during the study period, broken out by booster dose status.

The booster effectiveness estimate from the main GEE Poisson model is 56% (95% CI: [42%, 67%]). In addition to boosters, the incidence rate in our fitted GEE Poisson model depends on the student group and the date of completing the initial vaccine series **(Table 2)**. The incidence rate was significantly lower among students who completed the initial vaccine series after May 1, 2021 compared with those completing the initial vaccine series earlier. During the study period, the incidence rate was significantly higher among undergraduate students participating in fraternity and sorority activities (aIRR 2.16 [1.84, 2.54]) or belonging to athletic teams (aIRR 2.02 [1.57, 2.60]), and among professional students at the business school (aIRR 1.64 [1.15, 2.32]), when compared with other undergraduates. Students vaccinated with Ad26.COV2.S had a higher risk of infection relative to other vaccines, but difference was not statistically significant in our model (**Table 1, Supplementary Table S2**).

**Table 2:**
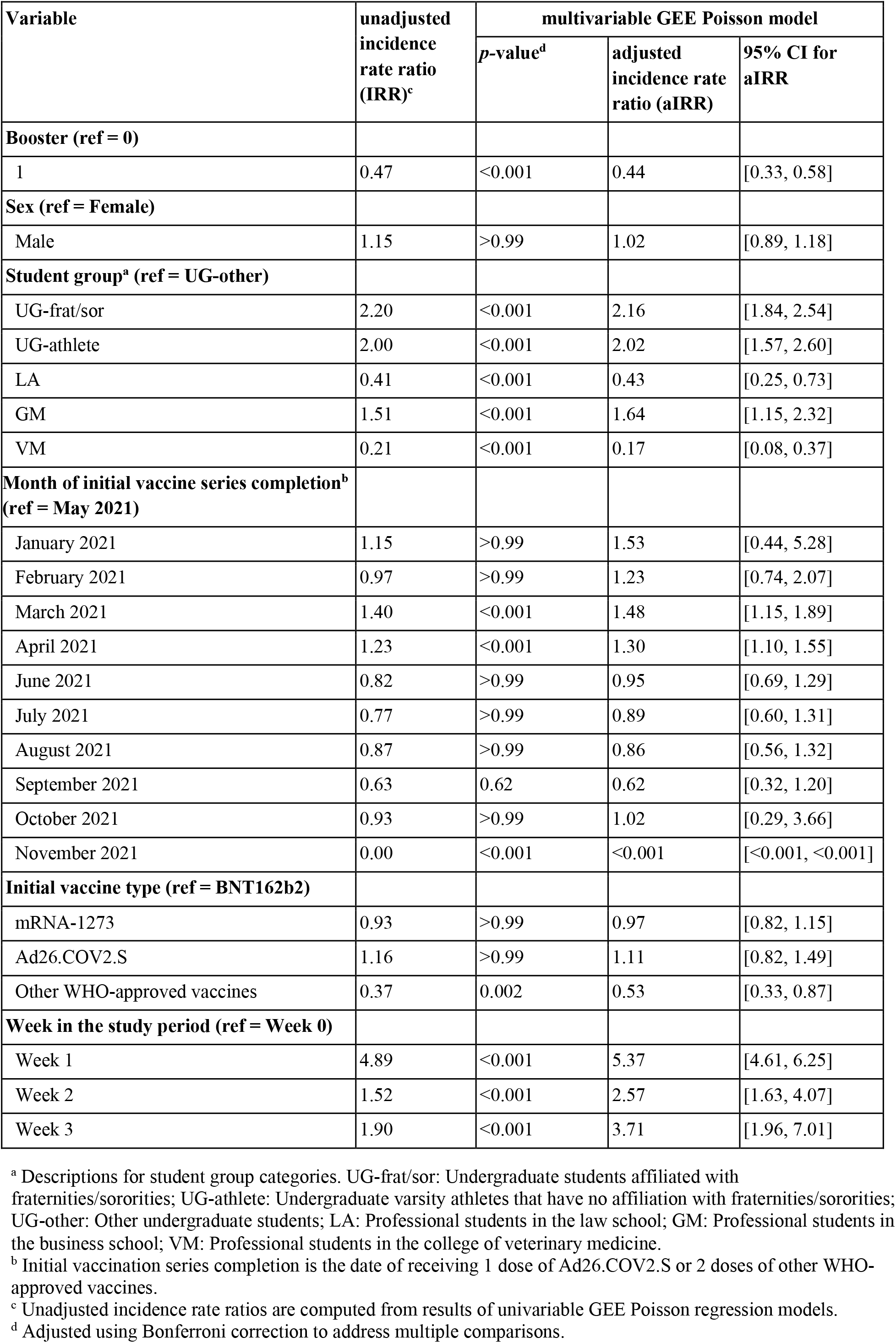
Summary of the full GEE Poisson regression model with covariates for sex, student group, initial vaccination series completion date, initial vaccine type, and week in the study period.

To study robustness to population size variation as students left campus at the end of the semester, we performed a GEE Poisson regression (**Supplementary Table S3**) for a shorter period (December 5, 2021 to December 18, 2021). This yields a similar booster effectiveness of 57% (95% CI: [42%, 68%], suggesting that our estimate of booster effectiveness from the full regression model is robust to the effect of students leaving campus at the end of the semester and the accordingly paused mandatory surveillance. As an additional robustness check, we performed a GEE Poisson regression (**Supplementary Table S4**) with multiple classes for the booster status of a person-day (unboosted, 0-6 days after booster administration, ≥ 7 days after booster dose administration). As expected, a booster status corresponding to 0-6 days after booster dose administration was not found to have a statistically significant effect on PCR-diagnosed infection (P > 0.99) relative to being unboosted.

We further performed a sensitivity analysis to assess the impact of the assumed delay between booster administration and booster effectiveness. Although 5,993 students received their booster dose during the study period (**Supplementary Figure S4**), 5,210 of them received it after leaving the active surveillance program at the end of the semester. These students did not contribute any person-days to the study after their last surveillance test, so they contributed only unboosted person-days. For the 783 students who received a booster dose while in active surveillance, we varied this delay from 1 day to 14 days and calculated the booster effectiveness against infection using the full model (**Supplementary Figures S2 and S5**). The estimated booster effectiveness is consistently above 48% and is relatively flat after 7 days suggesting that our estimate of booster effectiveness is robust to variation in the time required to mount an effective immune response after booster vaccination. Similarly, the incidence in the boosted population does not vary substantially as we change the assumed delay between booster administration and effectiveness (**Supplementary Table S5**).

In a separate sensitivity analysis using logistic regression (with GEE) (**Supplementary Table S6**), the estimated booster effectiveness is 57% (95% CI: [43%, 68%]). The estimate from the logistic regression (with GEE) is close to the estimate from the full GEE Poisson regression model.

In a subset analysis (**Supplementary Section S2**) on the undergraduate population living in on-campus housing, we did not observe a significant change in the estimated booster effectiveness on the subpopulation whether or not an additional covariate for housing building was included in the regression. This subset analysis justifies the use of our regression model without controlling for housing type on the full study population, given that housing information is only available for the on-campus undergraduates.

## Discussion

This study is one of the first community studies to quantify booster vaccine effectiveness from an actively surveilled population of young adults. Our study provides evidence that booster vaccinations significantly reduced infections in university settings during periods when the Omicron BA.1 variant was predominant, supporting the implementation of booster vaccination requirements to minimize community transmission.

In this retrospective analysis of SARS-CoV-2 tests performed at Cornell University’s Ithaca campus during a 27-day period when Omicron BA.1 was the predominant variant, the incidence of COVID-19 infections was approximately halved among participants vaccinated with a booster dose of a WHO-approved COVID-19 vaccine when compared with vaccinated participants without a booster dose. The calculated booster effectiveness was 56% (95% CI: [42%, 67%]), which is slightly lower than a previously reported effectiveness against symptomatic COVID-like illness in adults (66%; 95% CI: [64%, 68%]) in the same time period [17]. Our estimate is similar to the reduction in cumulative incidence of reported infections associated with booster vaccination during an Omicron wave in Los Angeles County [33] and consistent with a pooled estimate of booster effectiveness (47%; 95% CI: [19%, 65%]) from a meta-analysis of studies completed during periods of Omicron predominance across the globe [34]. Importantly, our analysis includes both symptomatic and asymptomatic infections because the student population was under active surveillance, whereas none of the studies included in the meta-analysis examined a population under active surveillance [7, 35-40]. Interestingly, between two studies on the effectiveness of a second booster dose (i.e., fourth vaccine dose) [41, 42], one that used data collected during active surveillance [42] also found lower effectiveness than one that did not [41]. Students vaccinated with Ad26.COV2.S had a higher risk of infection relative to other vaccines, similar to other studies [43], though the difference was not statistically significant, likely because a small number of students had Ad26.COV2.S initial doses.

We observed lower incidence rates among students with more recent initial vaccinations, consistent with waning protection against SARS-CoV-2 infection observed within a few months of completing the initial vaccination series [44]. Months in which fewer individuals completed their initial vaccine series did not have a statistically significant effect, consistent with fewer data points resulting in less statistical power. Varsity athletes and students in fraternities/sororities at institutions of higher education may have more social contact than other undergraduates; thus, consistent with previous studies, we found that this population may be at higher risk for SARS-CoV-2 infection [31, 45, 46]. As in other higher-education institutions [46], contact tracing at Cornell identified fraternity/sorority gatherings as significant spreading events. Local case investigation efforts also pointed to additional events, including post-Thanksgiving break travel and a series of end-of-semester gatherings, as contributing to Omicron spread. With the emergence of highly transmissible variants, travel and social gatherings may put students at increased risk of SARS-CoV-2 infection. We did not find a booster status corresponding to 0-6 days after booster dose administration to have a statistically significant effect on PCR-diagnosed infection relative to being unboosted. This could be because of an insufficient immune response to the booster within 6 days of administration, consistent with previous findings [22], or because of a lack of statistical power resulting from a small number of person days in the 0-6 day window after booster administration.

Our Poisson regression analysis is similar to Bar-On et al. [47], a previous study of vaccine booster effectiveness against COVID-19. See also [41] for another similar approach to estimating vaccine booster effectiveness against COVID-19. There are two common alternative statistical approaches to estimating vaccine effectiveness. The first approach is the emulated trial design (e.g., [48]), which matches subjects with the same covariates in an observational study to emulate a randomized trial. This design offers more robustness against misspecification of the functional form by which booster status and infection risk depend on covariates, but requires a larger dataset to enable matching simultaneously on all covariates.

We did not have a sufficiently large dataset to match on all the covariates included in our model. A second alternative approach is the test-negative case-control design [17], which controls bias by including those individuals who sought testing, and improvements that model the propensity to test [49]. Since testing was mandatory and compliance was high, it is not necessary to model the propensity to test in this study population.

## Limitations and Conclusion

We report on limitations of our approach that should be considered in interpreting our results. First, as we did not have an *a priori* protocol or statistical analysis plan (because the analysis was initially done for institutional research purposes), and because some analysis choices were made after observing the data (see **Supplementary Section S1**), confidence intervals and hypothesis tests would be invalidated by correlation that could have been unintentionally introduced between the data and the choice of analysis plan. Second, a limitation of the Poisson regression is information bias due to the fact that individuals were not tested every day. Delays in reporting a positive test would cause the period of the delay to have a misclassified infection status, whether this period is within one exposure period or spans multiple exposure periods. Third, there may be additional confounding bias because, even after controlling for covariates, people who avoid risk could also be more likely to get a booster vaccination, which could result in the boosted group having decreased exposure to SARS-CoV-2. This would cause our regression analyses to overestimate vaccine effectiveness. Conversely, if people engage in more risky behavior in response to perceived protection offered by booster vaccination, our analyses would underestimate vaccine effectiveness. Fourth, our data do not allow distinguishing between booster doses and additional doses for immunocompromised individuals, or distinguishing between symptomatic and asymptomatic infections for confirmed PCR-positive cases. As mentioned earlier, not all samples were tested for S-gene dropout and sequencing of every PCR-positive case was not performed. If boosters are more effective against the Delta variant than the Omicron variant, then Delta infections during the study period would lead our analysis to overestimate booster effectiveness against Omicron. Fifth, enough COVID-19 cases were observed to provide sufficient statistical power for moderately precise booster effectiveness estimates, but not to estimate how booster effectiveness varies with the manufacturer of the booster dose or the original vaccine. Most students were boosted with BNT162b2 in this study (**Supplementary Table S2**). Sixth, we did not include information on previous COVID-19 infections and had no information on the students’ underlying medical conditions or use of non-pharmaceutical interventions, although students were required to wear a mask (cloth, medical, or respirator) while inside campus buildings. This may lead to confounding bias if medical history differentially affected booster choice or if booster choice affected the use of non-pharmaceutical interventions. Seventh, gaps in time between a student’s last test and their departure from Ithaca (or the end of the study period, whichever came first) could lead to attrition bias. Eighth, although nearly 90% of positive samples tested for S-gene dropout during the study period exhibited this marker of the Omicron BA.1 variant [20], and sequencing confirmed the presence of the Omicron variant in all samples sequenced during this period, not all samples were tested for S-gene dropout and sequencing of each PCR-positive case was not performed. Finally, not every individual who was boosted before or during the study period may have uploaded their vaccination record, which may have led to information bias towards the null.

In conclusion, our findings are consistent with the notion that booster vaccine doses, relative to being vaccinated with the initial series, are effective in reducing infections in young adults during Omicron-predominant periods, thereby helping universities and other institutions to remain open safely. In the two months subsequent to the period covered in this study, almost the entire Cornell student body, if eligible, received at least one booster dose.

## Data Availability

Raw data used in this study cannot be shared publicly because they contain protected health information. Researchers who wish to request access to the data may contact Cornell's Office of the Vice President for Research and Innovation (contact), which oversees research access to Cornell COVID-19 surveillance data.

## Acknowledgments

We would like to thank the large number of people at Cornell University who supported the university’s response to COVID-19. JW and MO were supported by the Cornell University Office of the Provost. CC was supported by the David and Lucile Packard Foundation 2021-72608. SH was supported by NSF CMMI-2035086. MO was supported by the Atkinson Postdoctoral Fellowship. DS was supported by NSF DCCF-1740822 and NSF DCCF-1522054. PF was supported by AFOSR FA9550-19-1-0283.

## Supplementary Appendix

### Section S1: Analysis Timeline

In this section we describe how the analysis methodology appearing in this article was chosen and adapted during the reviewing process. This is provided to support understanding the potential for statistical bias from adjustments to the statistical analysis in response to the data.

Some of the authors (Wan, Frazier, Clarkberg, Henderson) first analyzed the data used in this paper in December 2021 and January 2022 as part of Cornell University’s institutional response to the pandemic. This analysis was done to understand the effect of booster vaccines to help plan interventions (e.g., the frequency of asymptomatic screening) for Cornell’s Spring 2022 semester and was made publicly available on Cornell’s website in early February 2022 [1].

The analysis began on December 17, 2021. Prompted by a hypothesis that booster doses would decrease the risk of infection, and that this would be important for planning for the spring 2022 semester, Clarkberg calculated population size and counts of infections diagnosed after December 5, 2021 broken out by (1) whether the individual had provided to Cornell by that date proof of receiving a booster dose; (2) whether that person was an undergraduate, graduate/professional student, or non-student employee. At this time, only 1,239 students had uploaded documentation of having received a booster dose, and only 2,055 employees had uploaded documentation, as many would not upload their documentation until closer to Cornell’s deadline of January 30, 2022. These counts showed that those who had uploaded documentation of receiving a booster dose had a smaller rate of diagnosed infection after December 5, 2021. It was understood that there was the potential for bias from confounding and because documentation of booster doses was missing for most individuals.

The data was broken out by student status (undergraduate, grad/professional, employee) because past data collected in the institutional response showed that infection rates were very different between these groups. The date of December 5, 2021 was chosen as the start of the study period because this date was understood within those working in Cornell’s institutional response to the pandemic to be the beginning of the Omicron surge.

These counts were recomputed on December 21, 2021, by which point 2,148 students and 2,262 employees had uploaded documentation of having received a booster. As they did on December 17, these counts showed that those who had uploaded documentation of receiving a booster shot had a smaller rate of diagnosed infection during the study period. A member of the institutional response (Dr. Lisa Nishii) asked to check for confounding due to fraternity and sorority members both having behaviors that put them more at risk of infection and having been more likely to receive a booster vaccine. Counts showed a similar effect for both fraternity/sorority members and other undergraduates.

At this point, starting on December 20, 2021 and continuing through late December 2021, Wan and Frazier planned the analysis presented in the report referenced above [1]. This planning was done without accessing the available data on which individuals had been boosted, except as described in the paragraph above. Indeed, the data on booster status available to the authors at this time was less than 17% of the data that would ultimately be collected (2,148 is 17% of the 13,065 students in the study population, and the count of 2,148 includes students who were not in the study population).

This analysis used a logistic regression on person days similar to the sensitivity analysis presented in the **Methods** section with the regression specification

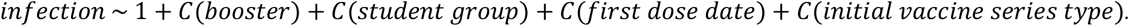

Compared to simple observations of counts, this analysis:

- Planned to use all of the available data on booster documentation. Most students analyzed in the analysis uploaded their documentation regarding booster vaccination after the analysis was planned, because Cornell announced a requirement after the study was planned requiring booster vaccinations with documentation provided by January 30, 2022.
- Focused only on students, because students had higher infection rates and so provided better statistical power and because they were the population of primary importance for public health planning with regard to the effect of booster vaccination on infection and its implications for surveillance testing (an important part of Cornell’s institutional response).
- Controlled for confounding using the student group covariate in the same way as we do elsewhere in this article. The use and definition of the student group covariate was chosen based on knowledge that this covariate explains a substantial amount of variation in infection risk at Cornell (see, e.g., [2, 3]).
- Controlled for confounding from the date and type of primary vaccination.
- Used logistic regression to control for multiple confounders and provide confidence intervals.

The analysis was identical to the logistic regression presented in the **Methods** section, except that:

- It assumed independent errors and used maximum likelihood estimation rather than assuming clustered errors and using generalized estimation equations.
- It did not control for the week when the student was diagnosed or the student’s sex.
- Rather than controlling for the date of primary vaccination using the month of completion of the last dose, as in the **Methods** section, it used a coarse discretization of when the first dose in the initial vaccine series was administered.
- Data on booster status was only available for students and employees who had uploaded their information at the time the analysis was posted. A substantial number of additional students uploaded records of having received a booster dose after this first analysis was completed to comply with Cornell’s decision to mandate boosters.

That report also included a sensitivity analysis, which was planned at the same time as the primary analysis (late December 2021), and which varied the assumed number of days between administration of a booster dose and when it is considered to become effective.

After this analysis was complete, we then planned our analysis for submission as a research article. For initial submission, we maintained a logistic regression over person days using independent errors and made the following modifications:

- We added sex as a covariate. We did this because one of the co-authors not associated with the original analysis (Lee) hypothesized that sex could cause confounding and because this is a common covariate controlled for in observational studies. This hypothesis was made without examining the data. We did not include sex as a covariate in our original analysis because experience with data from before the study period consistently suggested that sex is not predictive of infection risk among the student population at Cornell.
- We modified the covariate used to control for the date of vaccination to be a categorical variable indicating whether the date when a person was considered fully vaccinated (i.e., 14 days after completion of a primary vaccination series) was in (1) before May 2021 (23% of boosted individuals) (2) in May 2021 (49%) (3) in June 2021 (15%) or (4) after June 2021 (14%). We used this categorization because we were concerned about using a finer categorization for periods before May 2021 and after June 2021 when the fraction of the study population boosted in each month was small. Many students received booster doses in May or June when they first became widely available.

We also presented a sensitivity analysis using Poisson regression that used the same set of covariates. Each individual provided one datapoint if they were considered boosted during the whole study period, and 2 datapoints if they were considered unboosted at the beginning of the study period and boosted later on in the study period. Datapoints were modeled as independent.

Then, during the reviewing process and at the suggestion of reviewers and editors, we made the following changes to our main analysis:

- The Poisson regression, formerly a sensitivity analysis, became the main analysis. It was also changed to include a separate datapoint for each week, as discussed in the main paper. The logistic regression was retained for sensitivity analysis.
- We included the week during the study period as a covariate. This was added to control for confounding due to changes in infection risk over time, which could have been correlated with the fraction of the population boosted in that week.
- We added “other vaccines” in addition to the three initial vaccine series types we had considered previously (i.e., BNT162b2, mRNA-1273, and Ad26.COV2.S).
- We changed the covariate used to control for when the primary vaccine series was completed to include a categorical variable for each month. This was done to address a concern from a reviewer that the previous categorization was ad hoc.
- We moved from assuming independent errors to using generalized estimation equations to control for within-individual correlation in infection risk that could arise from behavior that is either persistently less risky or more risky than the general population.

We also conducted several sensitivity analyses to respond to questions from the reviewing team that are presented in the **Methods** section and **Supplementary Section S2**. We also conducted a sensitivity analysis including the manufacturer of the booster, which was not included in the article because we hypothesized before conducting the analysis that the sample size would be too small to estimate differences in booster effectiveness between vaccine manufacturers and because the analysis, once conducted, confirmed this hypothesis.

Estimates of vaccine booster effectiveness were not sensitive to these changes in the analysis. The analysis in the institutional report [1] estimated that a booster dose reduces the odds of having a PCR-detected SARS-CoV-2 infection relative to an initial vaccination series by 54% (95% confidence interval [45%, 62%], P <0.001). The analysis in the first submission as a research article estimated that a booster dose reduces the odds of infection by 52% (95% confidence interval [37%, 64%], P<0.001). For comparison, after incorporating comments from the reviewing team, our main analysis estimates that a booster dose further reduces the rate of infection relative to an initial vaccination series by 56% (95% confidence interval [42%, 67%], P <0.001).

### Section S2: Additional Subset Analysis

At the suggestion of the reviewers, to assess the robustness of our regression results to additional kinds of student clustering, we performed a subset analysis on the undergraduates living in on-campus housing, including the building in which they lived as an additional covariate.

We performed a GEE Poisson regression analysis (**Supplementary Table S1**) on the on-campus undergraduate population (*n* = 6,570) whose housing information is available. We added the housing building in which they lived at the beginning of the study period as an additional covariate in the regression model.

When the covariate for housing building was included, we obtained an estimated booster effectiveness of 53% (95% CI: [16%, 74%]) (**Supplementary Table S1**). This estimate remains close to the estimate from the full model using GEE Poisson regression (i.e., 56%), but has a wider confidence interval due to the reduced study population. All but one p-values associated with housing buildings were found statistically significant at the 0.05 level, indicating insufficient evidence of confounding in infections due to on-campus housing. (The *p*-values and confidence intervals for the covariates associated with housing buildings are concealed here due to institutional concern of being able to recover building names from their statistics.)

To assess the robustness of our subset analysis, we performed a second GEE Poisson regression (**Supplementary Table S1**) on the same on-campus undergraduate population, without controlling for the housing building. This yields an estimated booster effectiveness of 51% (95% CI: [18%, 71%]). We observe that whether or not housing building was included as an additional covariate did not significantly change the regression result on the same subpopulation. This justifies the use of our regression model without controlling for housing building on the full study population, given that housing information is only available for the on-campus undergraduates.

## Section S3: Supplementary Tables and Figures

**Supplementary Table S1:** Summary of the GEE Poisson regression model for the on-campus undergraduate population with and without housing building as an additional covariate, and other covariates for sex, student group, initial vaccine series completion date, initial vaccine type, and week during the study period.

| Variable | multivariable GEE Poisson model (on-campus undergraduate population) |  |  |  |  |  |
| --- | --- | --- | --- | --- | --- | --- |
|  | adjusting for housing building |  |  | without adjusting for housing building |  |  |
|  | *p*-value<sup>c</sup> | adjusted incidence rate ratio (aIRR) | 95% CI for aIRR | *p*-value<sup>c</sup> | adjusted incidence rate ratio (aIRR) | 95% CI for aIRR |
| **Booster (ref = 0)** |  |  |  |  |  |  |
| 1 | <0.001 | 0.47 | [0.26, 0.84] | <0.001 | 0.49 | [0.29, 0.82] |
| **Sex (ref = Female)** |  |  |  |  |  |  |
| Male | >0.99 | 1.03 | [0.80, 1.33] | >0.99 | 1.00 | [0.79, 1.26] |
| **Student group<sup>a</sup> (ref = UG-other)** |  |  |  |  |  |  |
| UG-frat/sor | <0.001 | 2.19 | [1.58, 3.03] | <0.001 | 2.27 | [1.70, 3.04] |
| UG-athlete | <0.001 | 2.18 | [1.30, 3.64] | <0.001 | 2.17 | [1.38, 3.40] |
| **Month of initial vaccine series completion<sup>b</sup> (ref = May 2021)** |  |  |  |  |  |  |
| January 2021 | >0.99 | 2.88 | [0.57, 14.58] | >0.99 | 2.60 | [0.52, 13.13] |
| February 2021 | >0.99 | 0.94 | [0.30, 2.92] | >0.99 | 0.91 | [0.32, 2.64] |
| March 2021 | >0.99 | 1.28 | [0.73, 2.25] | >0.99 | 1.29 | [0.77, 2.15] |
| April 2021 | >0.99 | 1.14 | [0.84, 1.55] | >0.99 | 1.15 | [0.86, 1.52] |
| June 2021 | >0.99 | 0.97 | [0.60, 1.58] | >0.99 | 0.94 | [0.60, 1.47] |
| July 2021 | >0.99 | 1.08 | [0.61, 1.92] | >0.99 | 1.07 | [0.63, 1.81] |
| August 2021 | >0.99 | 1.12 | [0.58, 2.14] | >0.99 | 1.13 | [0.62, 2.04] |
| September 2021 | >0.99 | 0.80 | [0.32, 2.00] | >0.99 | 0.81 | [0.35, 1.88] |
| October 2021 | >0.99 | 1.47 | [0.31, 7.09] | >0.99 | 1.39 | [0.31, 6.32] |
| November 2021 | <0.001 | <0.001 | [<0.001, <0.001] | <0.001 | <0.001 | [<0.001, <0.001] |
| **Initial vaccine type (ref = BNT162b2)** |  |  |  |  |  |  |
| mRNA-1273 | >0.99 | 0.88 | [0.64, 1.22] | 0.73 | 0.82 | [0.61, 1.09] |
| Ad26.COV2.S | >0.99 | 0.92 | [0.49, 1.71] | >0.99 | 0.88 | [0.50, 1.56] |
| Other WHO-approved vaccines | 0.950 | 0.54 | [0.23, 1.28] | 0.180 | 0.51 | [0.23, 1.11] |
| **Week in the study period (ref = Week 0)** |  |  |  |  |  |  |
| Week 1 | <0.001 | 6.22 | [4.71, 8.20] | <0.001 | 6.13 | [4.75, 7.92] |
| Week 2 | >0.99 | 1.43 | [0.20, 10.48] | >0.99 | 1.28 | [0.21, 8.03] |
| Week 3 | 0.36 | 4.39 | [0.72, 26.83] | 0.41 | 3.77 | [0.67, 21.27] |

<sup>a</sup> Descriptions for student group categories. UG-frat/sor: Undergraduate students affiliated with fraternities/sororities; UG-athlete: Undergraduate varsity athletes that have no affiliation with fraternities/sororities;
UG-other: Other undergraduate students; LA: Professional students in the law school; GM: Professional students in the business school; VM: Professional students in the college of veterinary medicine. <sup>b</sup> Initial vaccination series completion is the date of receiving 1 dose of Ad26.COV2.S or 2 doses of other WHO- approved vaccines. <sup>c</sup> Adjusted using Bonferroni correction to address multiple comparisons.

**Supplementary Table S2:** Distribution of booster dose type among students in the study population that received their booster dose before or during the study period, broken out by initial vaccination and booster dose type.

|  | **Initial vaccination type** |  |  |  |
| --- | --- | --- | --- | --- |
|  | BNT162b2 | mRNA-1273 | Ad26.COV2.S | Other vaccines |
|  | *n* (col %) | *n* (col %) | *n* (col %) | *n* (col %) |
| **Booster dose type** |  |  |  |  |
| BNT162b2 | 4,262 (42%) | 454 (11%) | 168 (21%) | 207 (29%) |
| mRNA-1273 | 878 (9%) | 1,644 (39%) | 129 (16%) | 84 (12%) |
| Ad26.COV2.S | 8 (0%) | 5 (0%) | 27 (3%) | 0 (0%) |
| Other vaccines<sup>a</sup> | 1 (0%) | 0 (0%) | 0 (0%) | 2 (0%) |
| Unboosted or boosted after the study period | 4,918 (49%) | 2,111 (50%) | 491 (60%) | 411 (58%) |
| **Total** | 10,067 (100%) | 4,214 (100%) | 815 (100%) | 704 (100%) |

<sup>a</sup> Other vaccines approved by WHO for emergency use.

**Supplementary Figure S1:**
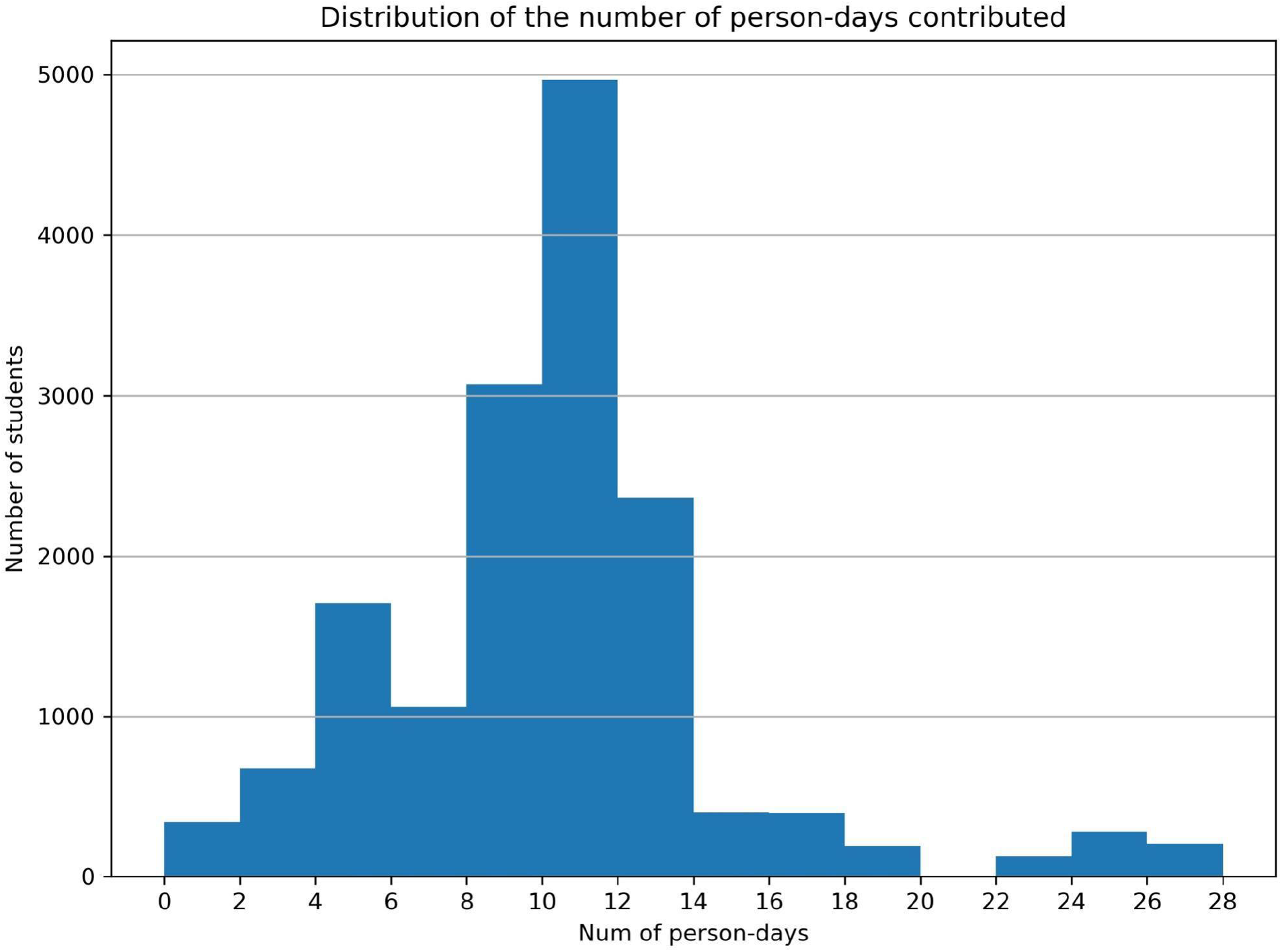
Distribution of the number of person-days contributed by each student in the study population (n = 15,800). Mean = 9.8 person-days; Median = 10 person-days; Interquartile range = 4 person-days.

**Supplementary Figure S2.**
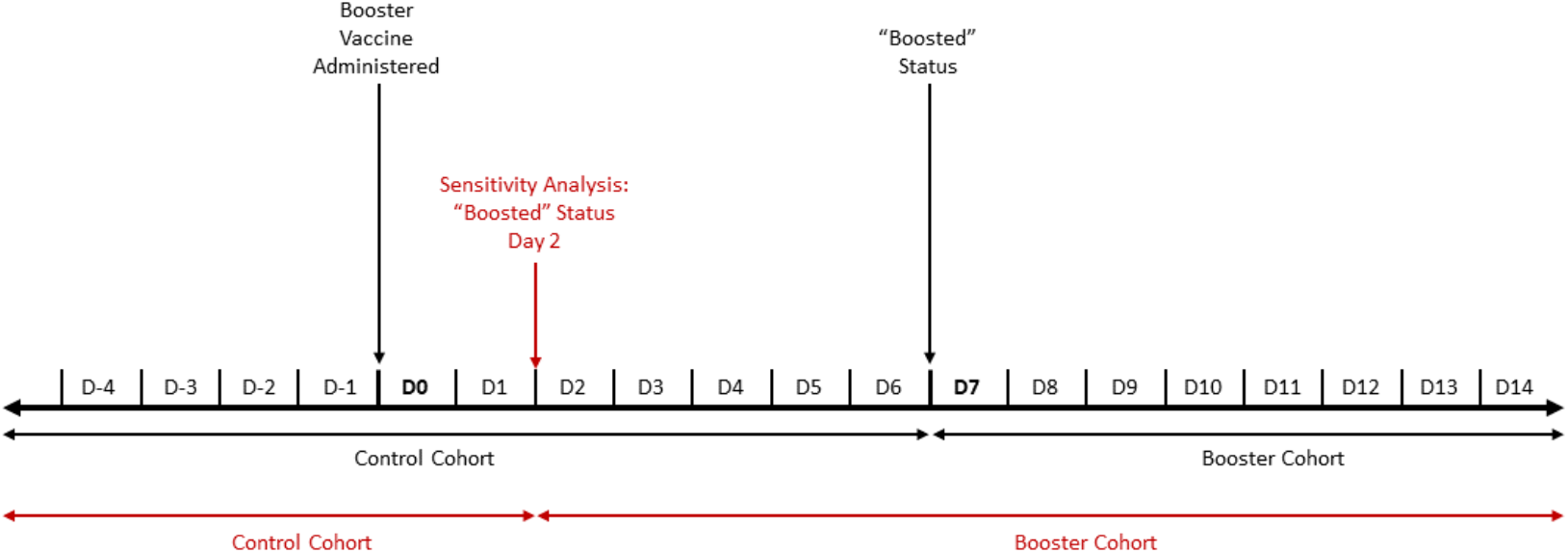
Allocation of person-days to the control and booster cohorts. The black timeline shows the main analysis, with students having a “boosted” status 7 days after SARS-CoV-2 booster vaccine administration. The red timeline shows one example of our sensitivity analysis varying the time to “boosted” status, with students in this example achieving a “boosted” status 2 days after booster vaccine administration.

**Supplementary Figure S3:**
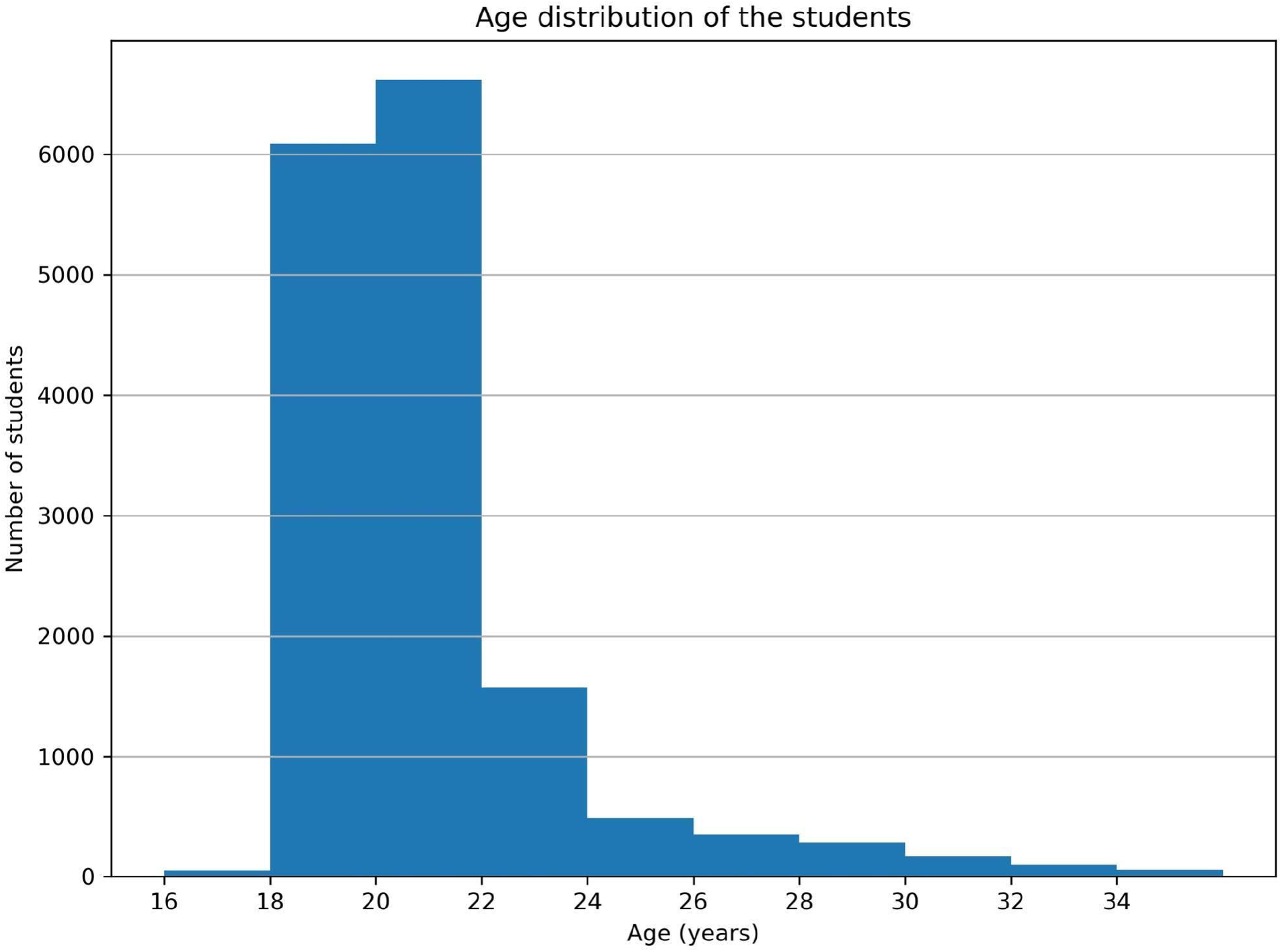
Age distribution of students in the study population (n =15,800). Age was estimated from birth year. Mean age = 20.6 years; Median age = 20 years; Interquartile range = 2 years.

**Supplementary Figure S4:**
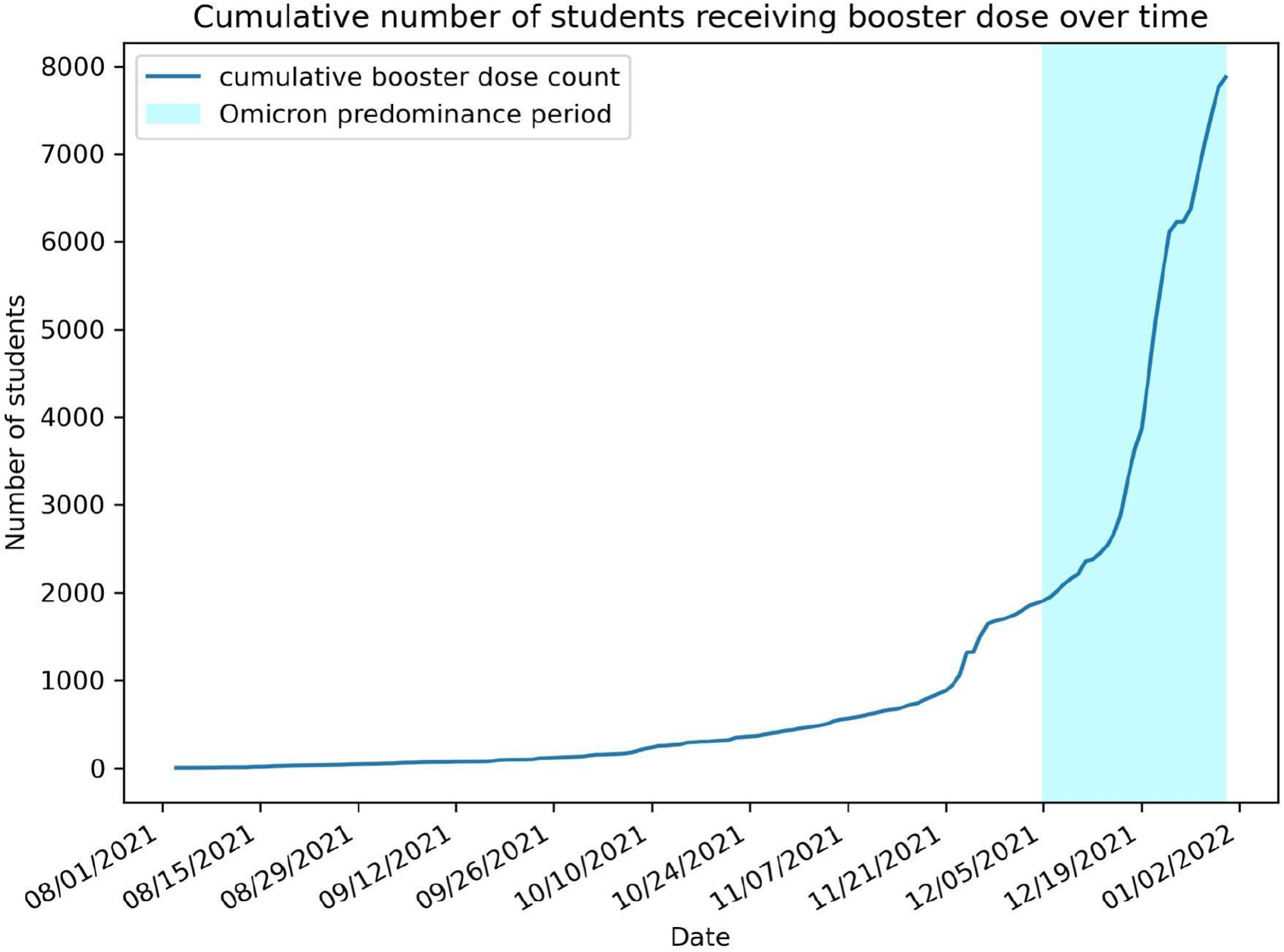
Cumulative number of students receiving COVID-19 booster dose, over time. The shaded region represents the Omicron predominance period.

**Supplementary Table S3:** Summary of the GEE Poisson regression model in the shortened study period (Dec 5, 2021 to Dec 18, 2021) with covariates for sex, student group, initial vaccine series completion date, initial vaccine type, and week during the study period.

| Variable | multivariable GEE Poisson model<br>(shortened study period) |  |  |
| --- | --- | --- | --- |
|  | *p*-value<sup>c</sup> | adjusted incidence rate ratio (aIRR) | 95% CI for IRR |
| **Booster (ref = 0)** |  |  |  |
| 1 | <0.001 | 0.43 | [0.32, 0.58] |
| **Sex (ref = Female)** |  |  |  |
| Male | >0.99 | 1.00 | [0.87, 1.16] |
| **Student group<sup>a</sup> (ref = UG-other)** |  |  |  |
| UG-frat/sor | <0.001 | 2.14 | [1.82, 2.51] |
| UG-athlete | <0.001 | 2.10 | [1.64, 2.70] |
| LA | <0.001 | 0.32 | [0.16, 0.62] |
| GM | <0.001 | 1.79 | [1.26, 2.55] |
| VM | <0.001 | 0.10 | [0.03, 0.32] |
| **Month of initial vaccine series completion<sup>b</sup> (ref = May 2021)** |  |  |  |
| January 2021 | >0.99 | 1.72 | [0.52, 5.70] |
| February 2021 | >0.99 | 1.27 | [0.76, 2.13] |
| March 2021 | <0.001 | 1.52 | [1.19, 1.95] |
| April 2021 | <0.001 | 1.30 | [1.09, 1.54] |
| June 2021 | >0.99 | 0.97 | [0.71, 1.32] |
| July 2021 | >0.99 | 0.88 | [0.59, 1.32] |
| August 2021 | >0.99 | 0.78 | [0.49, 1.25] |
| September 2021 | 0.37 | 0.55 | [0.26, 1.18] |
| October 2021 | >0.99 | 1.10 | [0.31, 3.84] |
| November 2021 | <0.001 | <0.001 | [<0.001, <0.001] |
| **Initial vaccine type (ref = BNT162b2)** |  |  |  |
| mRNA-1273 | >0.99 | 0.96 | [0.81, 1.14] |
| Ad26.COV2.S | >0.99 | 1.08 | [0.80, 1.46] |
| Other WHO-approved vaccines | 0.006 | 0.53 | [0.31, 0.90] |
| **Week in the study period (ref = Week 0)** |  |  |  |
| Week 1 | <0.001 | 5.38 | [4.62, 6.26] |

<sup>a</sup> Descriptions for student group categories. UG-frat/sor: Undergraduate students affiliated with fraternities/sororities; UG-athlete: Undergraduate varsity athletes that have no affiliation with fraternities/sororities; UG-other: Other undergraduate students; LA: Professional students in the law school; GM: Professional students in the business school; VM: Professional students in the college of veterinary medicine.
<sup>b</sup> Initial vaccination series completion is the date of receiving 1 dose of Ad26.COV2.S or 2 doses of other WHO-approved vaccines.
<sup>c</sup> Adjusted using Bonferroni correction to address multiple comparisons.

**Supplementary Table S4:** Summary of the GEE Poisson regression model with multi-class booster status (divided into unboosted, 0-6 days after booster dose administration, and ≥ 7 days after booster dose administration) and covariates for sex, student group, initial vaccine series completion date, initial vaccine type, and week during the study period.

| Variable | multivariable GEE Poisson model<br>(multi-class booster status) |  |  |
| --- | --- | --- | --- |
|  | *p*-value<sup>c</sup> | adjusted incidence rate ratio (aIRR) | 95% CI for aIRR |
| **Booster (ref = Unboosted)** |  |  |  |
| 0 - 6 days after booster dose administration | >0.99 | 0.84 | [0.56, 1.26] |
| $\geq 7$ days after booster dose administration | <0.001 | 0.43 | [0.32, 0.58] |
| **Sex (ref = Female)** |  |  |  |
| Male | >0.99 | 1.02 | [0.89, 1.18] |
| **Student group<sup>a</sup> (ref = UG-other)** |  |  |  |
| UG-frat/sor | <0.001 | 2.16 | [1.83, 2.54] |
| UG-athlete | <0.001 | 2.03 | [1.57, 2.61] |
| LA | <0.001 | 0.43 | [0.25, 0.73] |
| GM | <0.001 | 1.65 | [1.16, 2.34] |
| VM | <0.001 | 0.17 | [0.08, 0.37] |
| **Month of initial vaccine series completion<sup>b</sup> (ref = May 2021)** |  |  |  |
| January 2021 | >0.99 | 1.56 | [0.45, 5.43] |
| February 2021 | >0.99 | 1.24 | [0.74, 2.09] |
| March 2021 | <0.001 | 1.49 | [1.16, 1.91] |
| April 2021 | <0.001 | 1.31 | [1.10, 1.55] |
| June 2021 | >0.99 | 0.94 | [0.69, 1.29] |
| July 2021 | >0.99 | 0.88 | [0.60, 1.30] |
| August 2021 | >0.99 | 0.85 | [0.55, 1.32] |
| September 2021 | 0.59 | 0.62 | [0.32, 1.19] |
| October 2021 | >0.99 | 1.02 | [0.28, 3.66] |
| November 2021 | <0.001 | <0.001 | [<0.001, <0.001] |
| **Initial vaccine type (ref = BNT162b2)** |  |  |  |
| mRNA-1273 | >0.99 | 0.98 | [0.82, 1.16] |
| Ad26.COV2.S | >0.99 | 1.11 | [0.82, 1.50] |
| Other WHO-approved vaccines | 0.001 | 0.53 | [0.33, 0.87] |
| **Week in the study period (ref = Week 0)** |  |  |  |
| Week 1 | <0.001 | 5.39 | [4.62, 6.28] |
| Week 2 | <0.001 | 2.62 | [1.65, 4.16] |
| Week 3 | <0.001 | 3.77 | [1.99, 7.15] |

<sup>a</sup> Descriptions for student group categories. UG-frat/sor: Undergraduate students affiliated with fraternities/sororities; UG-athlete: Undergraduate varsity athletes that have no affiliation with fraternities/sororities; UG-other: Other undergraduate students; LA: Professional students in the law school; GM: Professional students in the business school; VM: Professional students in the college of veterinary medicine.
<sup>b</sup> Initial vaccination series completion is the date of receiving 1 dose of Ad26.COV2.S or 2 doses of other WHO- approved vaccines. <sup>c</sup> Adjusted using Bonferroni correction to address multiple comparisons.

**Supplementary Figure S5:**
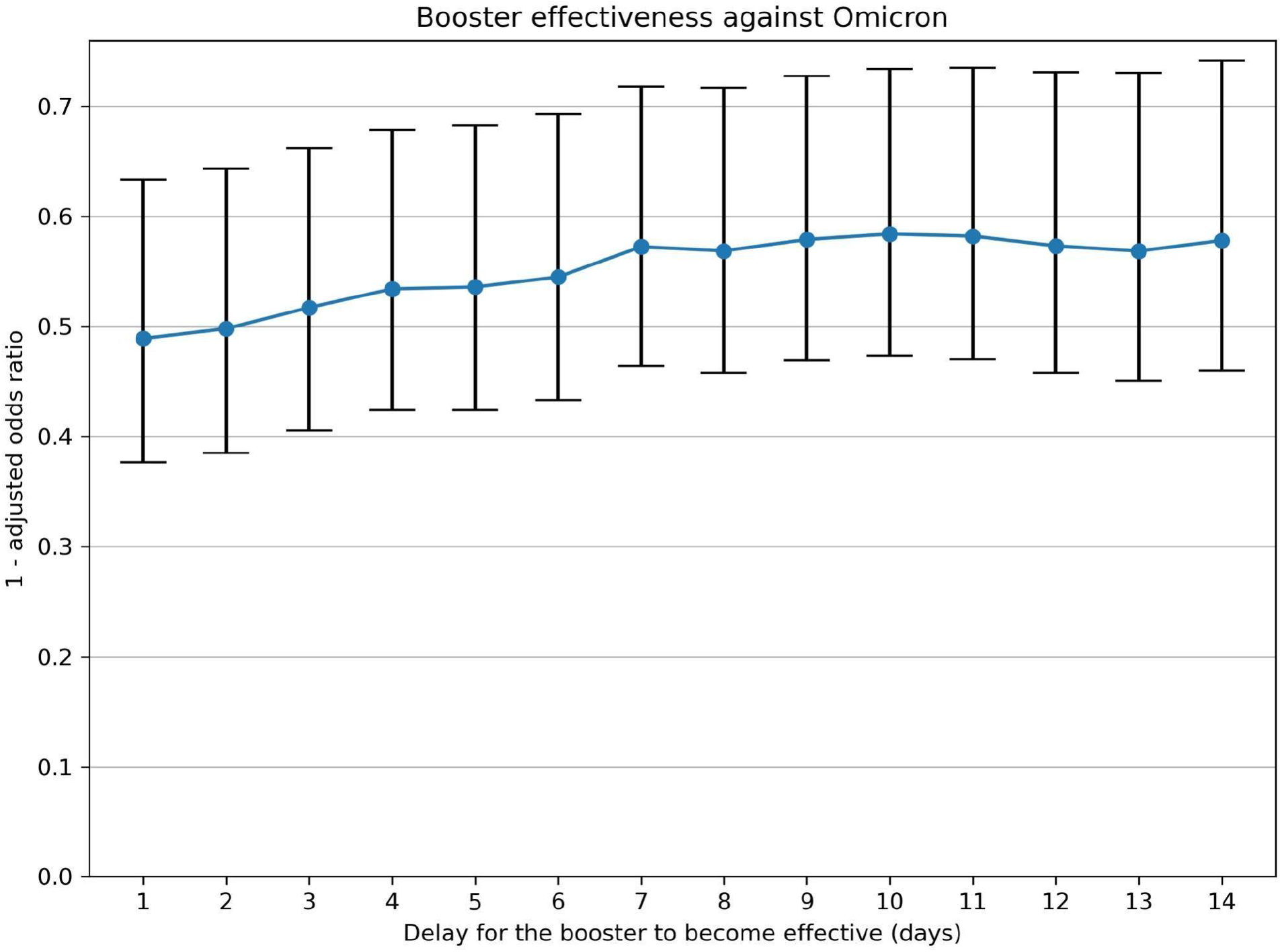
Mean and 95% confidence interval for the booster effectiveness against infection during the Omicron predominance period, as we vary the assumed delay for the booster dose to become effective after booster administration. Note that this analysis differs from our other sensitivity analysis considering the period shortly after booster administration: the analysis here combines all person days after the assumed delay into a single group, diluting the booster’s effect when the assumed delay is too short; the other sensitivity analysis separates people 0-6 days after booster administration from those that are 7+ days after.

**Supplementary Table S5:**
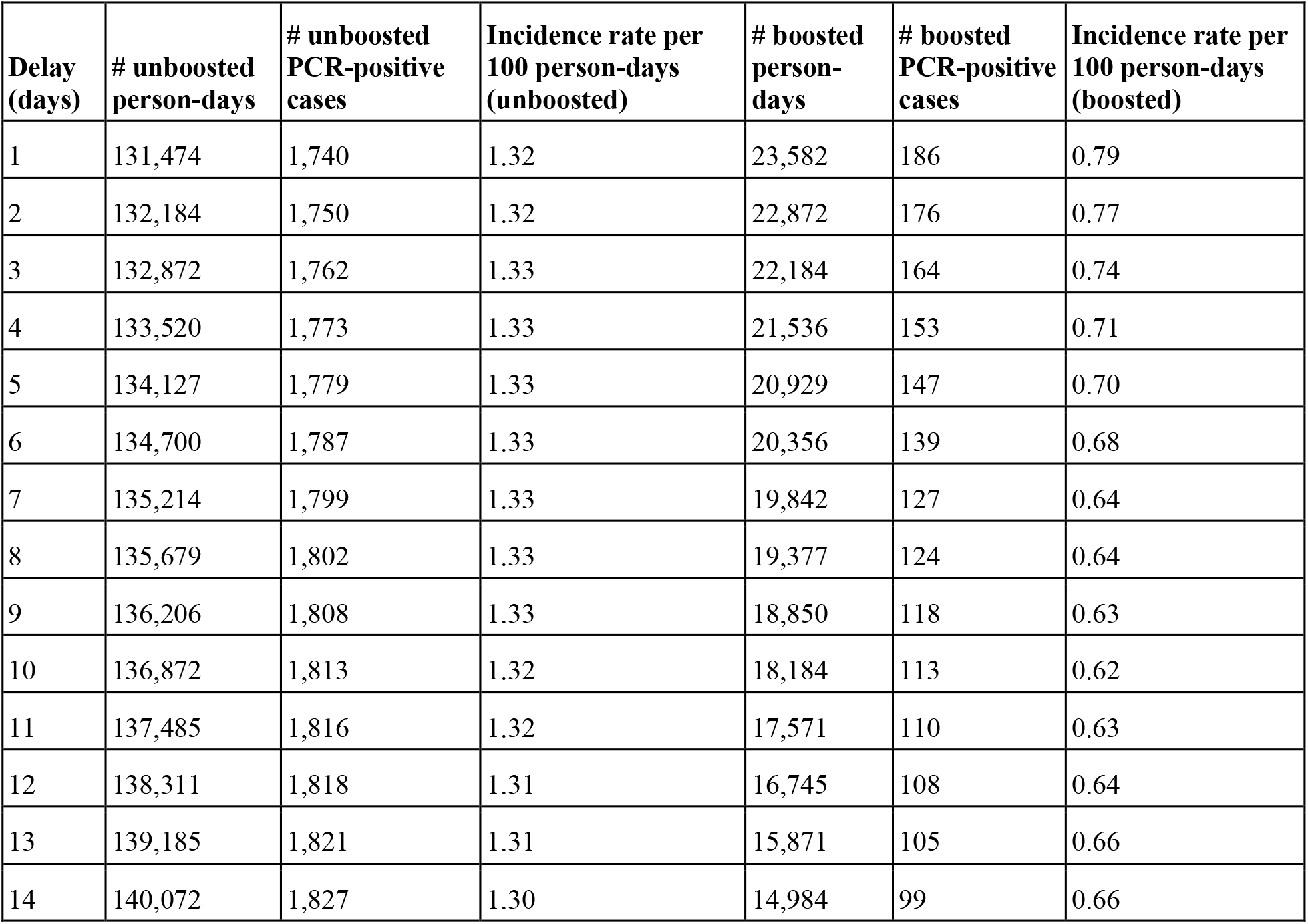
Number of unboosted and boosted person-days, PCR-positive cases, incidence rate with respect to different assumed delays for the booster to become effective after administration.

| **Delay (days)** | **# unboosted person-days** | **# unboosted PCR-positive cases** | **Incidence rate per 100 person-days (unboosted)** | **# boosted person-days** | **# boosted PCR-positive cases** | **Incidence rate per 100 person-days (boosted)** |
| --- | --- | --- | --- | --- | --- | --- |
| 1 | 131,474 | 1,740 | 1.32 | 23,582 | 186 | 0.79 |
| 2 | 132,184 | 1,750 | 1.32 | 22,872 | 176 | 0.77 |
| 3 | 132,872 | 1,762 | 1.33 | 22,184 | 164 | 0.74 |
| 4 | 133,520 | 1,773 | 1.33 | 21,536 | 153 | 0.71 |
| 5 | 134,127 | 1,779 | 1.33 | 20,929 | 147 | 0.70 |
| 6 | 134,700 | 1,787 | 1.33 | 20,356 | 139 | 0.68 |
| 7 | 135,214 | 1,799 | 1.33 | 19,842 | 127 | 0.64 |
| 8 | 135,679 | 1,802 | 1.33 | 19,377 | 124 | 0.64 |
| 9 | 136,206 | 1,808 | 1.33 | 18,850 | 118 | 0.63 |
| 10 | 136,872 | 1,813 | 1.32 | 18,184 | 113 | 0.62 |
| 11 | 137,485 | 1,816 | 1.32 | 17,571 | 110 | 0.63 |
| 12 | 138,311 | 1,818 | 1.31 | 16,745 | 108 | 0.64 |
| 13 | 139,185 | 1,821 | 1.31 | 15,871 | 105 | 0.66 |
| 14 | 140,072 | 1,827 | 1.30 | 14,984 | 99 | 0.66 |

**Supplementary Table S6:** Summary of the GEE logistic regression model with covariates for sex, student group, initial vaccine series completion date, initial vaccine type, and week during the study period.

| Variable | multivariable GEE logistic model |  |  |
| --- | --- | --- | --- |
|  | *p*-value<sup>c</sup> | adjusted odds ratio (aOR) | 95% CI for aOR |
| **Booster (ref = 0)** |  |  |  |
| 1 | <0.001 | 0.43 | [0.32, 0.57] |
| **Sex (ref = Female)** |  |  |  |
| Male | >0.99 | 1.02 | [0.89, 1.18] |
| **Student group<sup>a</sup> (ref = UG-other)** |  |  |  |
| UG-frat/sor | <0.001 | 2.21 | [1.87, 2.62] |
| UG-athlete | <0.001 | 2.07 | [1.59, 2.69] |
| LA | <0.001 | 0.42 | [0.25, 0.72] |
| GM | <0.001 | 1.66 | [1.16, 2.37] |
| VM | <0.001 | 0.16 | [0.07, 0.36] |
| **Month of initial vaccine series completion<sup>b</sup> (ref = May 2021)** |  |  |  |
| January 2021 | >0.99 | 1.55 | [0.43, 5.57] |
| February 2021 | >0.99 | 1.24 | [0.73, 2.11] |
| March 2021 | 0.001 | 1.49 | [1.16, 1.93] |
| April 2021 | <0.001 | 1.31 | [1.10, 1.57] |
| June 2021 | >0.99 | 0.95 | [0.69, 1.30] |
| July 2021 | >0.99 | 0.88 | [0.59, 1.31] |
| August 2021 | >0.99 | 0.85 | [0.55, 1.33] |
| September 2021 | 0.58 | 0.62 | [0.32, 1.19] |
| October 2021 | >0.99 | 1.03 | [0.28, 3.77] |
| November 2021 | <0.001 | <0.001 | [<0.001, <0.001] |
| **Initial vaccine type (ref = BNT162b2)** |  |  |  |
| mRNA-1273 | >0.99 | 0.97 | [0.82, 1.16] |
| Ad26.COVS2.S | >0.99 | 1.11 | [0.81, 1.51] |
| Other WHO-approved vaccines | 0.001 | 0.53 | [0.32, 0.86] |
| **Week in the study period (ref = Week 0)** |  |  |  |
| Week 1 | <0.001 | 5.58 | [4.77, 6.52] |
| Week 2 | <0.001 | 2.64 | [1.66, 4.19] |
| Week 3 | <0.001 | 3.88 | [2.04, 7.36] |

<sup>a</sup> Descriptions for student group categories. UG-frat/sor: Undergraduate students affiliated with fraternities/sororities; UG-athlete: Undergraduate varsity athletes that have no affiliation with fraternities/sororities; UG-other: Other undergraduate students; LA: Professional students in the law school; GM: Professional students in the business school; VM: Professional students in the college of veterinary medicine.
<sup>b</sup> Initial vaccination series completion is the date of receiving 1 dose of Ad26.COVS2.S or 2 doses of other WHO-approved vaccines.
<sup>c</sup> Adjusted using Bonferroni correction to address multiple comparisons.

## Notes

### Competing Interest Statement

The authors have declared no competing interest.

### Author Declarations

This work was completed as a part of Cornell's institutional planning and preparedness, designated as exempt from IRB review by the Cornell IRB.

### Summary of Updates

The manuscript has been revised to update the title, author affiliations, abstract, introduction section, methods section, discussion section, limitations and conclusion section, acknowledgements, and supplementary appendix.

